# Identifying 31 novel breast cancer susceptibility loci using data from genome-wide association studies conducted in Asian and European women

**DOI:** 10.1101/19003855

**Authors:** Xiang Shu, Jirong Long, Qiuyin Cai, Sun-Seog Kweon, Ji-Yeob Choi, Michiaki Kubo, Sue K. ParK, Manjeet K. Bolla, Joe Dennis, Qin Wang, Yaohua Yang, Jiajun Shi, Xingyi Guo, Bingshan Li, Ran Tao, Kristan J. Aronson, Kelvin Y.K. Chan, Tsun L. Chan, Yu-Tang Gao, Mikael Hartman, Weang Kee Ho, Hidemi Ito, Motoki Iwasaki, Hiroji Iwata, Esther M. John, Yoshio Kasuga, Ui Soon Khoo, Mi-Kyung Kim, Allison W. Kurian, Ava Kwong, Jingmei Li, Artitaya Lophatananon, Siew-Kee Low, Shivaani Mariapun, Koichi Matsuda, Keitaro Matsuo, Kenneth Muir, Dong-Young Noh, Boyoung Park, Min-Ho Park, Chen-Yang Shen, Min-Ho Shin, John J. Spinelli, Atsushi Takahashi, Chiuchen Tseng, Shoichiro Tsugane, Anna H. Wu, Yong-Bing Xiang, Taiki Yamaji, Ying Zheng, Roger L. Milne, Alison M. Dunning, Paul D.P. Pharoah, Montserrat García-Closas, Soo-Hwang Teo, Xiao-ou Shu, Daehee Kang, Douglas F. Easton, Jacques Simard, Wei Zheng

## Abstract

Common genetic variants in 183 loci have been identified in relation to breast cancer risk in genome-wide association studies (GWAS). These risk variants combined explain only a relatively small proportion of breast cancer heritability, particularly in Asian populations. To search for additional genetic susceptibility loci for breast cancer, we performed a meta-analysis of data from GWAS conducted in Asians (24,206 cases and 24,775 controls). Variants showing an association with breast cancer risk at *P* < 0.01 were evaluated in GWAS conducted in European women including 122,977 cases and 105,974 controls. In the combined analysis of data from both Asian and European women, the lead variant in 28 loci not previously reported showed an association with breast cancer risk at *P* < 5 ×10^−8^. In the meta-analysis of all GWAS data from Asian and European descendants, we identified SNPs in three additional loci in association with breast cancer risk at *P* < 5 ×10^−8^. The associations for 10 of these loci were replicated in an independent sample of 16,787 cases and 16,680 controls of Asian women (*P* < 0.05). Expression quantitative trait locus (eQTL) and gene-based analyses provided evidence for the possible involvement of the *YBEY, MAN2C1, SNUPN, TBX1, SEMA4A, STC1, MUTYH, LOXL2*, and *LINC00886* genes underlying the associations observed in eight of these 28 newly identified risk loci. In addition, we replicated the association for 78 of the 166 previously reported risk variants at *P* < 0.05 in women of Asian descent using GWAS data. These findings improve our understanding of breast cancer genetics and etiology and extend to Asian populations previous findings from studies of European women.

## INTRODUCTION

Breast cancer is the most commonly diagnosed malignancy and the leading cause of cancer-related deaths in women worldwide^1^. Genetic linkage studies and family-based studies have identified many high- and moderate-penetrance mutations in breast cancer predisposition genes, including *BRCA1, BRCA2, PTEN, ATM, PALB2*, and *CHEK2*^2^. In addition, large-scale genome-wide association studies (GWAS), conducted primarily in Asian and European women, have identified more than 180 susceptibility loci for breast cancer risk^3-8^. These identified loci explain a relatively small proportion of familial relative risk of breast cancer^8^.

The Asia Breast Cancer Consortium (ABCC) is the largest breast cancer GWAS consortium conducted in Asian-ancestry populations. We have shown previously that GWAS conducted in Asians could uncover cancer genetic risk variants that are either unique to the Asian population or more difficult to identify in studies conducted in European women^3,4,9-16^. It also has been shown that a large proportion of common susceptibility loci are shared between Asian and European populations, although the lead variants in many loci may differ between these two populations^6,8^. To search for novel breast cancer susceptibility loci, we conducted Asian-specific and cross-ancestry meta-analyses combining the data of the ABCC and the Breast Cancer Association Consortium (BCAC) with a total sample size of approximately 310,000 women (∼82,000 Asians and ∼228,000 Europeans). We herein report the discovery of 31 novel risk loci for breast cancer and the replication of a large number of known breast cancer susceptibility loci in Asian women.

## RESULTS

### Overall associations for newly associated loci

We identified 28 loci with at least one common variant at each locus showing a significant association with breast cancer risk in the cross-ancestry meta-analysis (i.e., *P* < 5 ×10^−8^) (Table 1 and Figure 1-a). None of these lead risk variants reside within a 500Kb region flanked by any of the 183 previously reported breast cancer risk variants. No obvious inflation in statistical estimates was observed for either Asian-specific or cross-ancestry meta-analysis after excluding known loci (sample size-adjusted λ _1000_ = 1.012 and 1.001, respectively; Supplementary Figure 1). No evidence of heterogeneity in associations was observed between the two racial populations except for rs2758598 and rs142360995 (Table 1, *P* _heterogeneity_ < 0.05, consistent in direction). The OR estimates for these 28 SNPs by study within the ABCC and BCAC consortia are presented in Supplementary Tables 2 and 3. We explored pleiotropic effects by assessing the newly identified lead variants and their correlated SNPs (in LD with r^2^ > 0.4 in either Asians or Europeans) from the online catalog of published GWAS (GWAS catalog). Pleiotropy was found for seven of the 28 newly-associated SNPs (Supplementary Table 4).

**Table 1.**
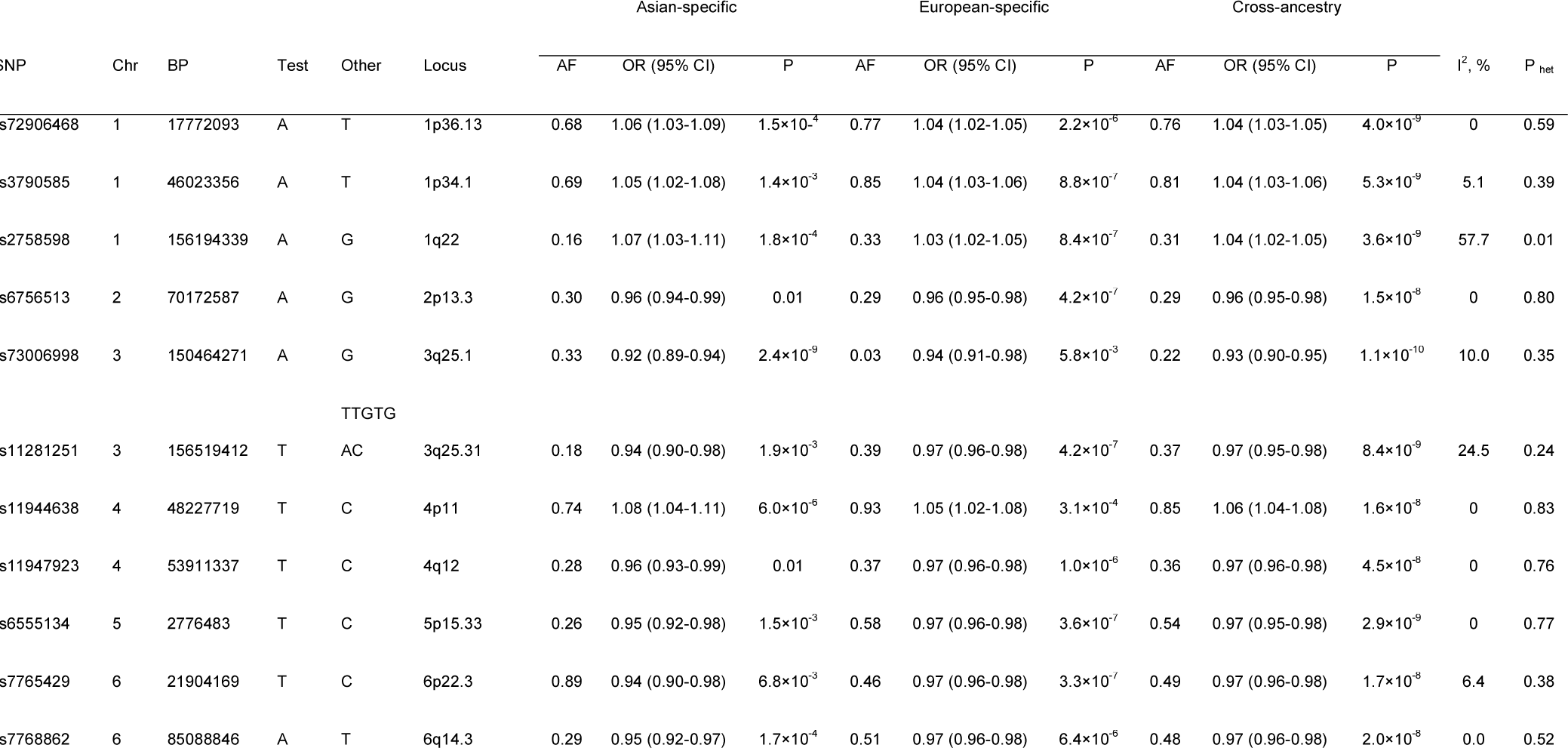
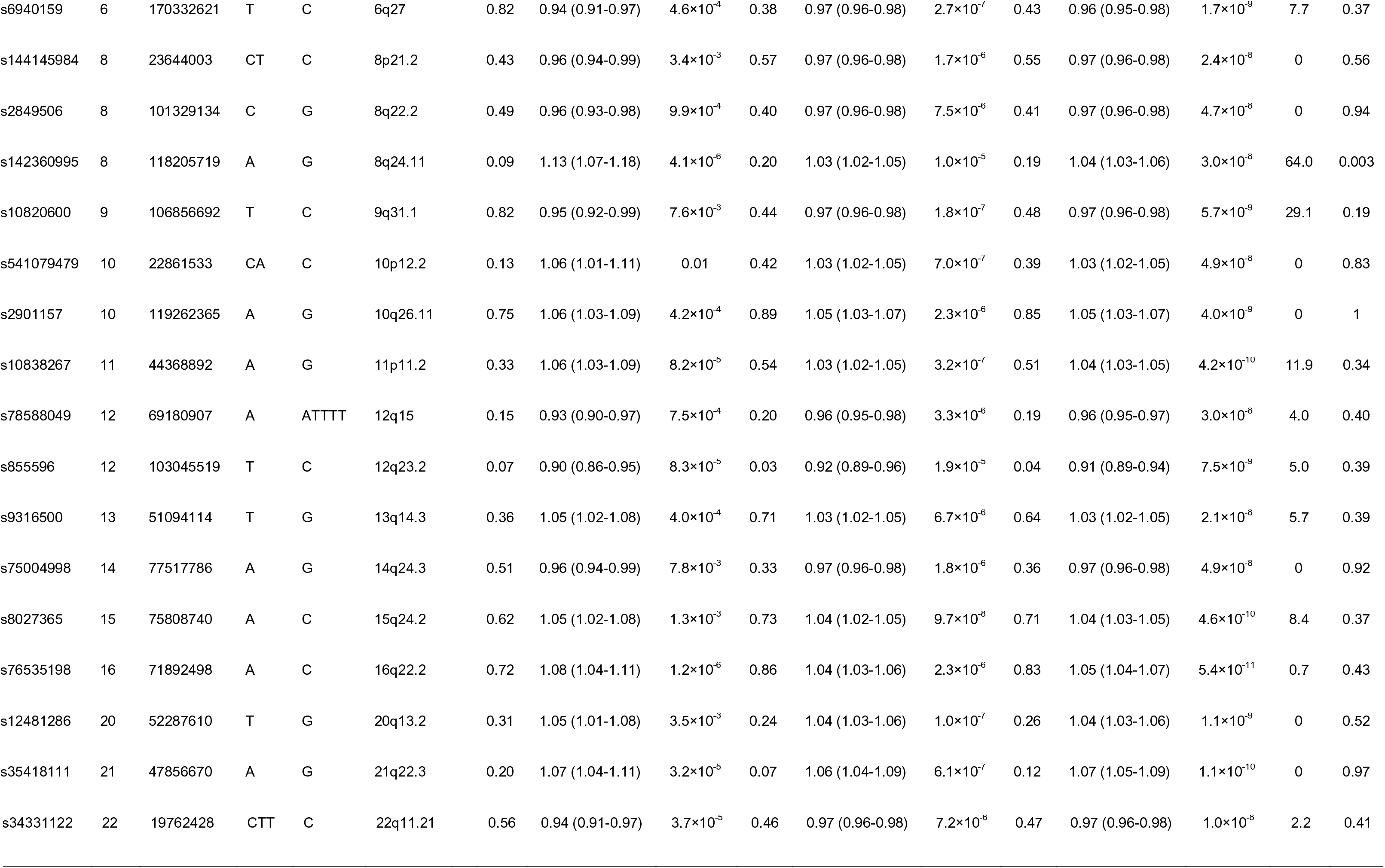
Twenty eight novel loci identified by the cross-ancestry Meta-analysis

**Table 2.** Association analysis of 28 newly-associated SNPs by estrogen receptor status

| SNP | Chr | BP | Test | Other | ER positive | | | ER negative | | | $I^2$, % | $P_{het}$ |
| --- | --- | --- | --- | --- | --- | --- | --- | --- | --- | --- | --- | --- |
|  |  |  |  |  | AF | OR (95% CI) | P | AF | OR (95% CI) | P |  |  |
| rs72906468 | 1 | 17772093 | A | T | 0.76 | 1.03 (1.02-1.05) | $6.9 \times 10^{-5}$ | 0.75 | 1.04 (1.01-1.06) | $2.0 \times 10^{-3}$ | 0 | 0.75 |
| rs3790585 | 1 | 46023356 | A | T | 0.81 | 1.05 (1.03-1.06) | $7.3 \times 10^{-7}$ | 0.80 | 1.03 (1.00-1.05) | 0.05 | 28.8 | 0.24 |
| rs2758598 | 1 | 156194339 | A | G | 0.32 | 1.03 (1.02-1.05) | $3.6 \times 10^{-5}$ | 0.31 | 1.02 (1.00-1.04) | 0.10 | 0 | 0.37 |
| rs6756513 | 2 | 70172587 | A | G | 0.29 | 0.97 (0.95-0.98) | $6.9 \times 10^{-6}$ | 0.29 | 0.98 (0.96-1.00) | 0.12 | 33.4 | 0.22 |
| rs73006998 | 3 | 150464271 | A | G | 0.20 | 0.91 (0.88-0.93) | $3.6 \times 10^{-10}$ | 0.24 | 0.96 (0.92-1.00) | 0.07 | 81.5 | 0.02 |
| rs11281251 | 3 | 156519412 | T | TTGTGAC | 0.37 | 0.96 (0.95-0.98) | $1.7 \times 10^{-7}$ | 0.36 | 0.96 (0.94-0.98) | $3.9 \times 10^{-4}$ | 24.5 | 0.24 |
| rs11944638 | 4 | 48227719 | T | C | 0.88 | 1.07 (1.04-1.09) | $8.5 \times 10^{-7}$ | 0.86 | 1.03 (1.00-1.07) | 0.07 | 45.8 | 0.17 |
| rs11947923 | 4 | 53911337 | T | C | 0.36 | 0.97 (0.95-0.98) | $2.4 \times 10^{-6}$ | 0.36 | 0.96 (0.94-0.98) | $4.5 \times 10^{-4}$ | 0 | 0.79 |
| rs6555134 | 5 | 2776483 | T | C | 0.55 | 0.96 (0.95-0.98) | $1.4 \times 10^{-7}$ | 0.53 | 0.97 (0.95-0.99) | $8.4 \times 10^{-3}$ | 0 | 0.45 |
| rs7765429 | 6 | 21904169 | T | C | 0.49 | 0.96 (0.94-0.97) | $8.8 \times 10^{-10}$ | 0.50 | 1.00 (0.98-1.02) | 0.79 | 90.1 | 0.002 |
| rs7768862 | 6 | 85088846 | A | T | 0.48 | 0.97 (0.96-0.98) | $1.6 \times 10^{-5}$ | 0.47 | 0.97 (0.95-0.99) | $2.6 \times 10^{-3}$ | 0 | 0.92 |
| rs6940159 | 6 | 170332621 | T | C | 0.42 | 0.97 (0.95-0.98) | $1.8 \times 10^{-6}$ | 0.44 | 0.97 (0.95-1.00) | 0.02 | 0 | 0.49 |
| rs144145984 | 8 | 23644003 | CT | C | 0.55 | 0.96 (0.95-0.97) | $1.3 \times 10^{-8}$ | 0.54 | 1.00 (0.97-1.02) | 0.65 | 86.9 | 0.006 |
| rs2849506 | 8 | 101329134 | C | G | 0.41 | 0.97 (0.95-0.98) | $1.5 \times 10^{-6}$ | 0.42 | 0.99 (0.97-1.01) | 0.15 | 55.5 | 0.13 |
| rs142360995 | 8 | 118205719 | A | G | 0.20 | 1.04 (1.02-1.06) | $4.0 \times 10^{-6}$ | 0.19 | 1.04 (1.01-1.06) | $7.4 \times 10^{-3}$ | 0 | 0.72 |
| rs10820600 | 9 | 106856692 | T | C | 0.48 | 0.97 (0.96-0.99) | $2.9 \times 10^{-4}$ | 0.49 | 0.96 (0.94-0.98) | $4.5 \times 10^{-4}$ | 0 | 0.36 |
| rs541079479 | 10 | 22861533 | CA | C | 0.40 | 1.04 (1.02-1.05) | $1.0 \times 10^{-6}$ | 0.38 | 1.03 (1.00-1.05) | 0.02 | 0 | 0.45 |
| rs2901157 | 10 | 119262365 | A | G | 0.86 | 1.05 (1.02-1.07) | $2.8 \times 10^{-5}$ | 0.85 | 1.05 (1.02-1.08) | $1.5 \times 10^{-3}$ | 0 | 0.81 |
| rs10838267 | 11 | 44368892 | A | G | 0.52 | 1.03 (1.02-1.05) | $9.4 \times 10^{-6}$ | 0.51 | 1.04 (1.01-1.06) | $7.9 \times 10^{-4}$ | 0 | 0.75 |
| rs78588049 | 12 | 69180907 | A | ATTTT | 0.19 | 0.95 (0.93-0.97) | $3.1 \times 10^{-9}$ | 0.19 | 0.98 (0.96-1.01) | 0.21 | 79.7 | 0.03 |
| rs855596 | 12 | 103045519 | T | C | 0.04 | 0.92 (0.88-0.95) | $3.9 \times 10^{-6}$ | 0.05 | 0.93 (0.88-0.98) | $5.4 \times 10^{-3}$ | 0 | 0.74 |
| rs9316500 | 13 | 51094114 | T | G | 0.65 | 1.03 (1.02-1.05) | $2.4 \times 10^{-5}$ | 0.63 | 1.02 (1.00-1.04) | 0.11 | 8.3 | 0.30 |
| rs75004998 | 14 | 77517786 | A | G | 0.36 | 0.97 (0.96-0.98) | $2.2 \times 10^{-5}$ | 0.37 | 0.97 (0.95-0.99) | $3.2 \times 10^{-3}$ | 0 | 0.96 |
| rs8027365 | 15 | 75808740 | A | C | 0.71 | 1.04 (1.02-1.05) | $8.0 \times 10^{-7}$ | 0.71 | 1.05 (1.03-1.08) | $9.9 \times 10^{-6}$ | 0 | 0.38 |
| rs76535198 | 16 | 71892498 | A | C | 0.83 | 1.05 (1.03-1.07) | $3.1 \times 10^{-6}$ | 0.83 | 1.06 (1.03-1.09) | $8.3 \times 10^{-5}$ | 0 | 0.55 |
| rs12481286 | 20 | 52287610 | T | G | 0.25 | 1.06 (1.04-1.07) | $6.9 \times 10^{-11}$ | 0.26 | 1.02 (0.99-1.04) | 0.20 | 85.1 | 0.01 |
| rs35418111 | 21 | 47856670 | A | G | 0.11 | 1.07 (1.04-1.09) | $6.8 \times 10^{-8}$ | 0.12 | 1.05 (1.02-1.09) | $3.6 \times 10^{-3}$ | 0 | 0.48 |
| rs34331122 | 22 | 19762428 | CTT | C | 0.47 | 0.96 (0.95-0.97) | $1.7 \times 10^{-8}$ | 0.48 | 0.98 (0.96-1.00) | 0.06 | 59.2 | 0.12 |

**Table 3.**
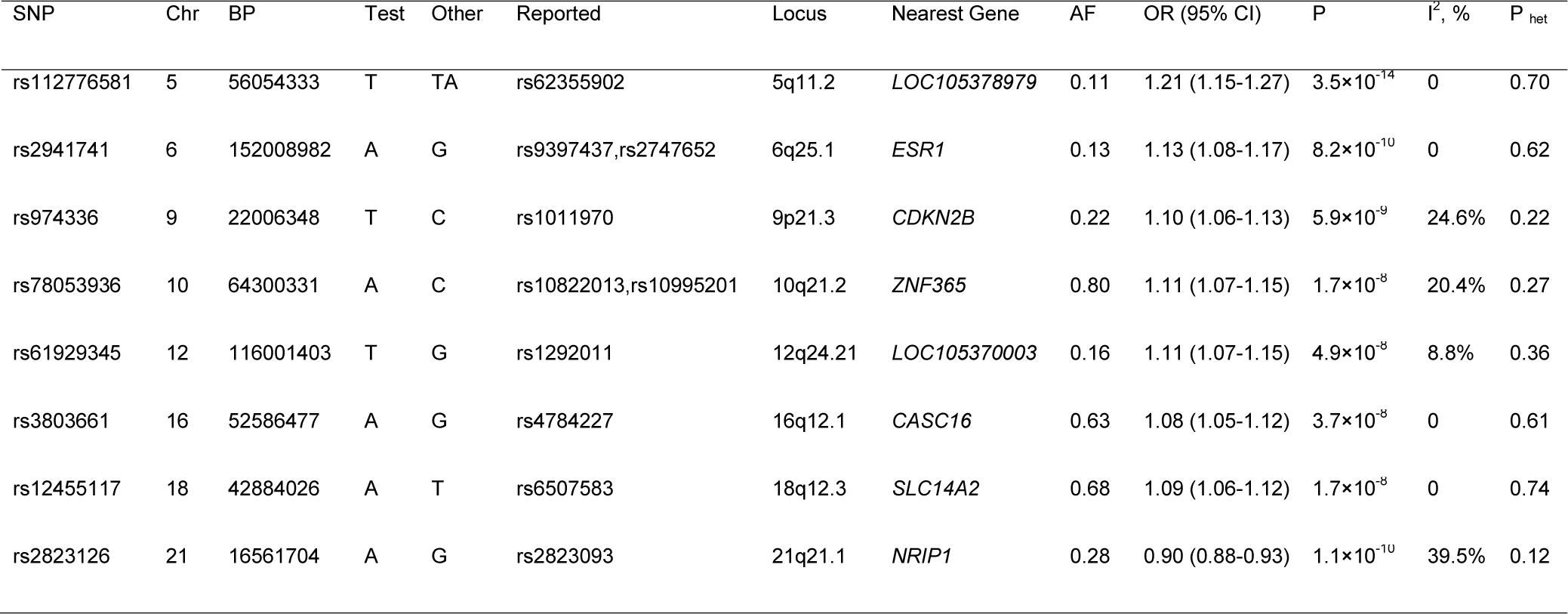
Eight novel breast cancer risk-associated SNPs located within previously known loci in Asians: A conditional analysis

**Figure 1. a.**
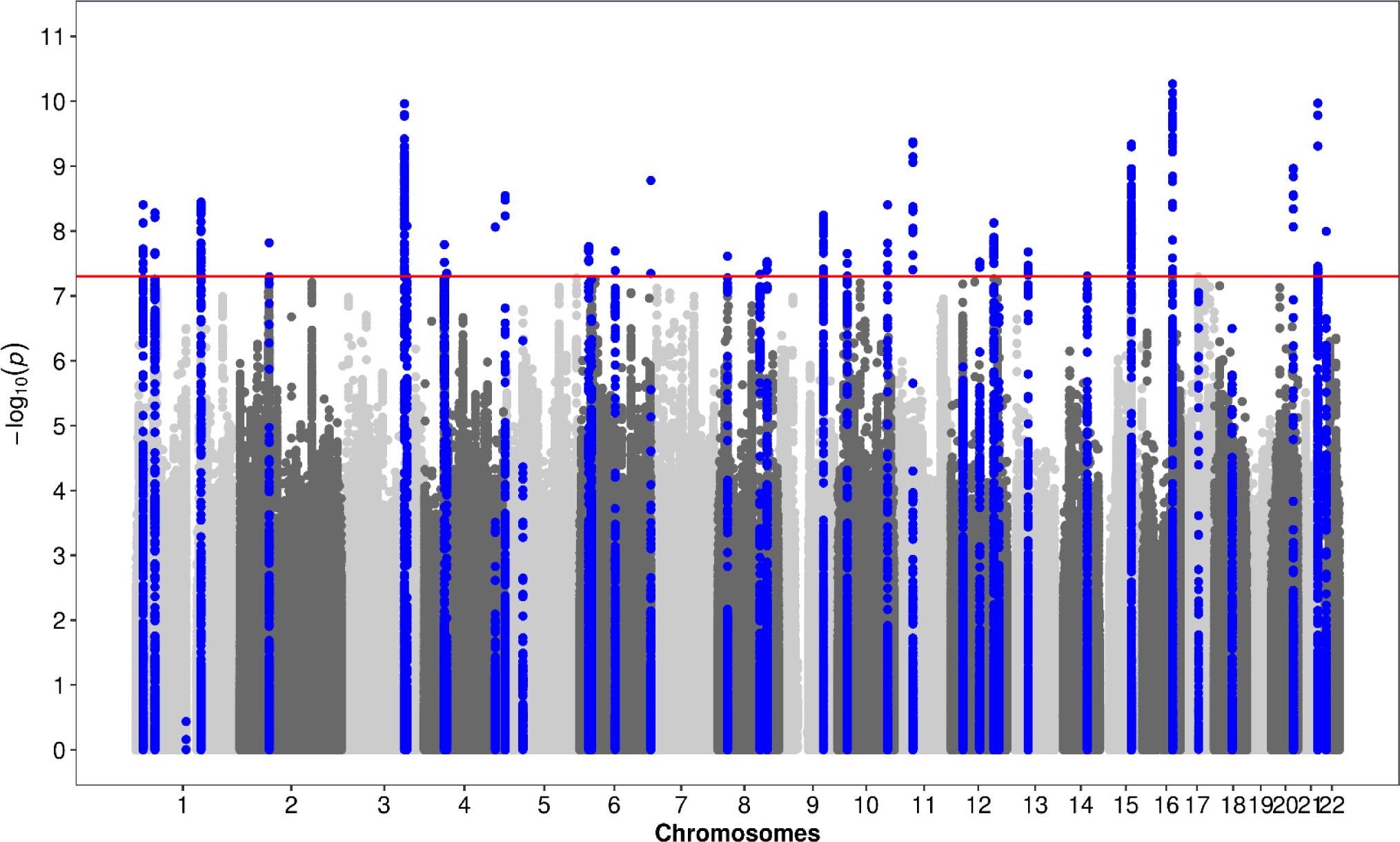
Manhattan plot of 28 newly-associated breast cancer susceptibility loci.

**Figure 1. b.**
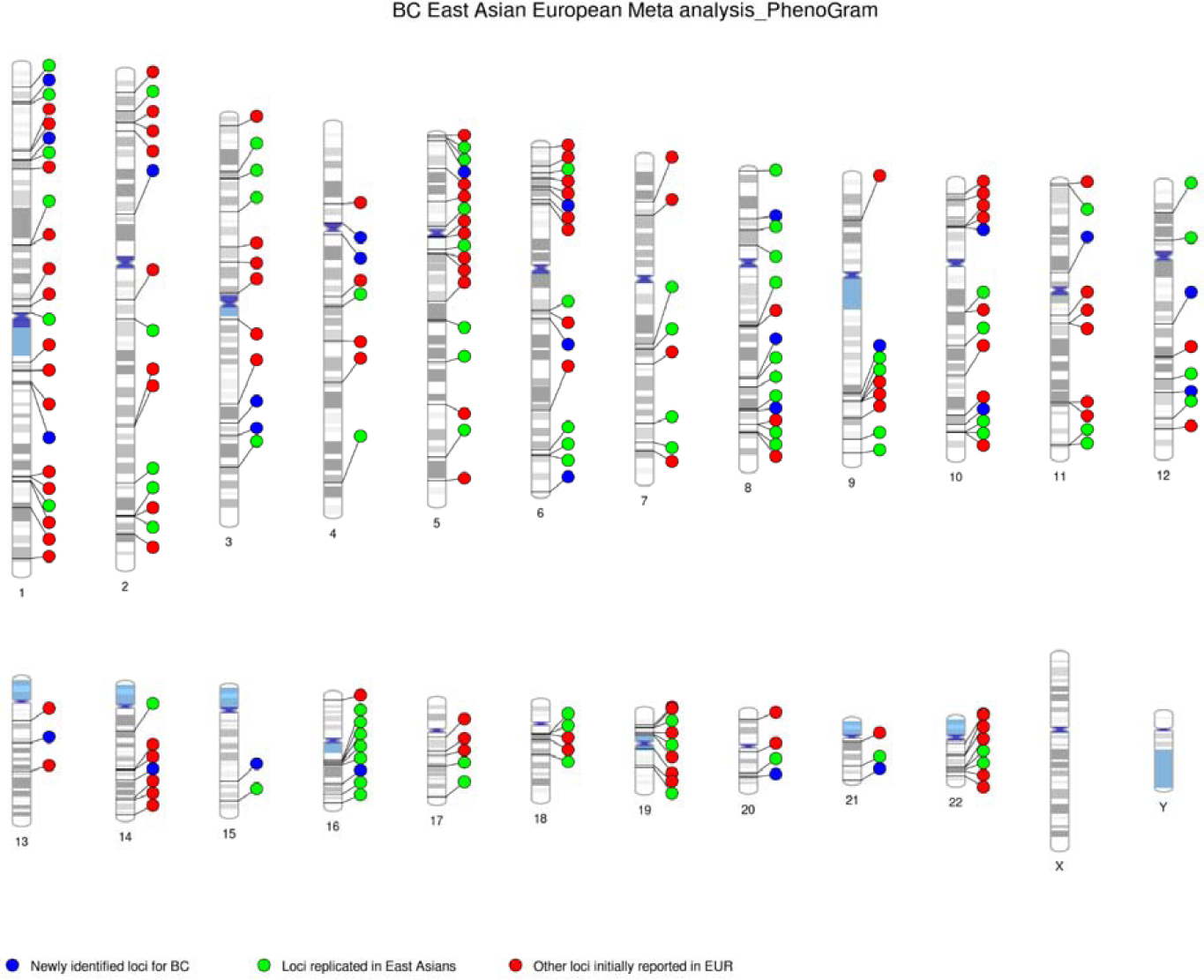
All breast cancer susceptibility loci identified to date. Blue: 28 newly-identified loci in the current study; Green: loci replicated at P Asian<0.05 in the current study; Red: loci not replicated in Asians or cannot be evaluated in the current study

All of the 28 SNPs showed a nominally significant association (*P* < 0.05) with ER-positive breast cancer risk (Table 2). Fourteen of the 28 risk SNPs were also associated with ER-negative breast cancer risk in the cross-ancestry meta-analysis (*P* < 0.05). Heterogeneity between ER+ and ER-breast cancer risk (*P* _heterogeneity_ < 0.05) was observed for rs73006998, rs7765429, rs144145984, rs78588049, and rs12481286.

Of the 28 SNPs, 22 were investigated in an independent set of 10,829 cases and 10,996 controls included in ABCC and an additional 5,958 cases and 5,684 controls from studies conducted in Malaysia and Singapore (see Methods). A significant association at *P*< 0.05 was found for 10 SNPs, all with the association direction consistent with our main findings (Supplementary Table 5). Among them, five SNPs showed significant associations at *P* < 2.3×10^−3^ (0.05/22), including rs3790585 (1p34.1), rs73006998 (3q25.1), rs6940159 (6q27), rs855596 (12q23.2), and rs75004998 (14q24.3).

To uncover possible secondary association signals in newly identified breast cancer susceptibility loci, we performed analyses for SNPs within flanking 500kb of each lead SNP, with adjustment for the lead SNPs within each dataset. We then conduced meta-analyses to combine the results across studies of Asian women. Six potential secondary associations were identified (conditional P < 1×10^−4^), and all correlated (r^2^> 0.1 in 1000 Genome East Asians) except for rs7693779, at 4p12 (Supplementary Table 6).

Of the 28 SNPs newly identified to be associated with breast cancer risk, 13 SNPs are intronic, one in UTR, and 14 in intergenic regions. Using data from ENCODE and Roadmap, we found that the majority of these 28 overlapped with genomic functional biofeatures that were indicative of promoters or enhancers (Supplementary Tables 7 and 8). Of the 28 lead SNPs, four (rs3790585 at 1p34.1, rs6756513 at 2p13.3, rs10820600 at 9q31.1, and rs78588049 at 12q15) intersected with chromosomal segments annotated as strong enhancers or active promoters in breast tissue-originated cell lines. When all SNPs that were in LD with the lead SNPs with r^2^> 0.8 in either Asians or Europeans were evaluated, evidence of regulatory function was found for an additional seven (i.e. 1q22-rs2758598, 3q25.1-rs73006998, 3q25.31-rs11281251, 8q22.2-rs2849506, 14q24.3-rs75004998, 15q24.2-rs8027365, and 21q22.3-rs35418111).

### eQTL and gene-based analyses

To identify target genes of the 28 newly identified lead SNPs, we conducted *cis*-eQTL analyses in four independent datasets in breast tissue. Nine eQTL associations were identified with a *P* < 0.05 with same association direction in two or more independent sets (Supplementary Table 9). Potential candidate genes identified in this analysis included *LINC00886, ybeY metallopeptidase* (*YBEY*), *snurportin 1* (*SNUPN*), *mannosidase alpha class 2C member 1* (*MAN2C1*), *T-Box 1* (*TBX1*), *MutY DNA glycosylase* (*MUTYH*), *lysyl oxidase like 2* (*LOXL2*), *stanniocalcin 1* (*STC1*), and *semaphorin 4A* (*SEMA4A*). SNP rs144145984 was the eQTL for both *LOXL2* and *STC1* genes, but the association for *STC1* is much stronger. Similarly, SNP rs8027365 was associated with expression levels of two genes, *MAN2C1* and *SNUPN*.

With the exception of *TBX1* and *LOXL2*, we were able to build breast-tissue and/or cross-tissue models for all other eQTL-identified candidate genes with a prediction R^2^ > 0.01 (Supplementary Table 10). Expressions of *LINC00886, YBEY, MAN2C1* and *SEMA4A* could be predicted with a high accuracy by both breast tissue and cross tissue models (R^2^ > 0.09). We imputed expressions of seven genes other than *TBX1* and *LOXL2* and showed that these genes were associated with breast cancer risk in either the ABCC or BCAC data at *P* < 0.05 (Supplementary Table 10). Of these, genes hypothesized to have a tumor-suppressor function included *LINC00886, MAN2C1, SNUPN*, and *STC1*, while *YBEY, SEMA4A*, and *MUTYH* may have an oncogenic role in breast carcinogenesis based on their associations with breast cancer risk (Supplementary Table 11)

### Associations of previously reported risk variants in Asians

Of the 183 risk variants of breast cancer reported previously, 11 and 172 were originally discovered in studies conducted in Asians and European-ancestry populations, respectively. We were able to investigate 166 variants because 15 variants originally discovered in European populations were (nearly) monomorphic in Asians and two in high LD with rs2747652 (*ESR1*, 6q25.1) were removed. Of the 166 SNPs, 78 were found to be associated with breast cancer risk at *P* < 0.05, while 131 showed associations that were consistent in direction with those originally reported (Supplementary Table 12). Associations for five variants achieved genome-wide significance (*P* < 5 ×10^−8^, Asians), with two at 6q25.1 (*ESR1* and *TAB2*), and one each at 15q26.1 (*PRC1*), 16q12.1 (*TOX3*), and 21q22.12 (*LINC00160*). Additionally, borderline genome-wide significant associations were found in seven loci including 2q14.1, 2q35, 3p24.1, 5q33.3, 9q33.3, 12p13.1 and 17q22 (*P* < 1 ×10^−6^ in Asians).

### Independent association signals within previously reported susceptibility loci of breast cancer

We searched extensively for additional independent associations in Asians by conducting conditional analysis for variants located 500kb of the 166 previously reported SNPs. A total of 820 SNPs from 21 loci were associated with breast cancer risk after conditioning on known risk variants in Asians (Supplementary Table 13). Eight loci, 5q11.2, 6q25.1, 9p21.3, 10q21.2, 12q24.21, 16q12.1, 18q12.3 and 21q21.1, may harbor independent association signals with genome-wide significance (Table 3, conditional *P* < 5 ×10^−8^ in Asians). Five of these eight loci, including 5q11.2, 9p21.3, 12q24.21, 18q12.3, and 21q21.1, have not previously been linked to breast cancer risk in Asian populations. Significant heterogeneity between Asian and European-ancestry populations was observed (*P* _heterogeneity_ < 0.05) at 5q11.2, 9p21.3, 12q24.21, 16q12.1, and 21q21.1, and the strength of the association was stronger in Asian than European-ancestry women.

## DISCUSSION

This large-scale meta-analysis, including approximately 310,000 women of Asian and European ancestry and represents the largest GWAS to identify genetic determinants for breast cancer. In addition to identifying 28 novel risk loci for breast cancer, we replicated in Asian women 78 of the GWAS-identified risk variants for breast cancer. Since the risk variants initially reported in European populations might not be the lead SNPs in Asians, we performed further analyses to show that 21 known susceptibility loci may harbor additional independent signals, of which 16 showed at least one stronger association than the originally reported risk SNP. Our study has generated substantial novel information to improve the understanding of breast cancer genetics and etiology and provides clues for future studies to functionally characterize the risk variants and candidate genes identified in our study.

Similar to other GWAS, nearly all of the newly identified risk variants mapped to intergenic regions or introns of genes. One exception was rs10820600, which is located in the 5’-UTR region of the *SMC2* gene. *SMC2* encodes the structural maintenance of chromosomes protein-2, an essential subunit of the condensin complex I and II. The protein is critically involved in chromosome condensation and segregation during cell cycles^17^. Emerging evidence shows that *SMC2* mutations and dysregulated expression are associated with multiple cancers^18^.

Of the thirteen lead risk variants located in the introns of genes, six showed strong evidence of cis-regulation for seven genes nearby, including *YBEY, SNUPN, MAN2C1, LINC00886, TBX1, SEMA4A, and MUTYH*. For example, the locus at 21q22.3 (rs35418111) showed compelling evidence of influencing expression of *YBEY*, a gene that encodes a highly conserved metalloprotein. Our gene-based analysis indicated a potential oncogenic role of *YBEY* in breast cancer development. Although the function of *YBEY* has not been fully elucidated, dysregulation of its expressions caused by copy number variation has been found in familial and early-onset breast cancer^19^, as well as colorectal cancer^20^. Further, we showed that *MAN2C1* may play a protective role against breast carcinogenesis in the gene-based analysis. However, another study found that *MAN2C1* promotes cancer growth via a negative regulation of phosphatase and tensin homolog (*PTEN*) function in prostate and breast cancer cell lines^21^. These results suggested that *MAN2C1* may have distinct functional impact on cancer initiation compared to that on tumor progression. Few studies have investigated the mechanistic roles of *LINC00886, SNUPN* and *SEMA4A* in cancer initiation. Germline mutations in *SEMA4A* have been linked to the predisposition of familial colorectal cancer type X^22^. Our study provides the first evidence linking these two genes to breast cancer susceptibility.

Potential candidate genes were also revealed by the newly associated variants lying in the intergenic regions between coding genes. *LOXL2 and STC1* were pinpointed as cis targets of rs144145984 at 8p21.2. LOXL2 is a member of the lysyl oxidase family of amine oxidases and STC1 belongs to the glycoprotein hormones family. Research regarding the functions of *LOXL2* and *STC1* in cancer development is limited. However, pre-clinical studies have implicated *LOXL2* and *STC1* in the progression of breast cancer^23,24^. Inhibiting *LOXL2* activity shows a 55-75% decrease in primary tumor volume in female athymic nude mice, which were implanted with MDA-MB-231 human breast cancer cells^23^. The reduction in tumor burden was suspected to be mediated by the inhibition of angiogenesis. A recent study suggested the role of *STC1* played in the breast tumorigenesis could be subtype-dependent^24^. A cancer promoting function was found in murine mammary tumor cells and human triple negative breast cancer lines (MDA-MB-231), while an opposite function was shown in luminal breast cancer lines (ER+/PR+, T47D cells).

The pleiotropy of rs855596 at 12q23.2 provided a plausible mechanistic link for the observed genetic association with breast cancer risk. The minor (T) allele of rs855596 is associated with decreased breast cancer risk and is linked to the minor allele G of the nearby rs703556 (r^2^ = 0.94 in EA and 0.43 in East Asians). The G allele of rs703556 is associated with lower mammographic dense area in women^25^. Mammographic density, an established risk factor for breast cancer^26^, is a measure based on the radiographic appearance of the breast by mammography. Several loci were related to other cancers or benign tumors. SNPs in 22q11.21, 1q22 and 4q12 were found to be associated with risk of prostate cancer^27^, testicular germ cell tumor^28^ and leiomyoma, respectively^29^. We hypothesize potential underlying mechanisms via hormone metabolism for these loci. Variants at 10p12.2 (*PIP4K2A*) indicated an association with risk of acute lymphoblastic leukemia^30^ and 6p22.3 (*CASC15*) with endometrial cancer^31^, lung cancer^32^, and neuroblastoma^33^. These regions implicated in genetic susceptibility across different types of cancers may serve as prioritized target of interest for future fine-mapping studies.

Notable racial heterogeneity was found for the loci at 1q22 (rs2758598) and 8q24.11 (rs142360995), which may reflect the differential regional LD structures and allele frequency between the two populations at these loci. The effect sizes in Asians are larger than those in European populations for both SNPs, over four times for rs142360995 and two times for rs2758598. The association at 3q25.1 (rs73006998) was dominant by estimates in Asians (ABCC: 2.4×10^−9^; in BCAC, *P*= 5.8×10^−3^), although no heterogeneity was observed. Previously, the same locus was reported to be associated with hormonal receptor-positive breast cancer, with a borderline genome-wide significance in a Japanese population (rs6788895, LD r^2^ = 0.76 in East Asians)^34^. We found significant heterogeneity by ER status for this locus and the association was primarily driven by ER-positive cancer. Racial heterogeneity was also observed for many known risk variants initially reported in European populations. It may be attributable to multiple factors including the Winner’s curse^35^, population-specific LD structure, and false positives in the original GWAS.

In summary, in this large GWAS including 147,183 breast cancer cases and 130,749 unaffected controls, we identified 31 novel breast cancer susceptibility loci by meta-analyzing data of two large consortia conducted in Asian and European women. Using an independent set of 16,787 cases and 16,680 controls of Asian ancestry, we replicated ten of these loci. As many of the associations were driven by GWAS of European women, the low replication rate is not unexpected. Unfortunately, we could not include a further independent dataset of European ancestry to optimize the power in the replication stage. Nevertheless, our study reveals many novel loci and potential targeted genes that may influence breast cancer susceptibility, although the possibility of false-positives cannot be completely ruled out. Future investigations are warranted to replicate our findings.

## METHODS

### Study population

The overall cross-ancestry meta-analysis was conducted using data from two large consortia, the ABCC and BCAC. Detailed descriptions of participating studies are included in Supplementary Text 1 File. Briefly, in the ABCC, genome-wide SNP data were generated from 24,206 breast cancer cases and 24,775 unaffected controls recruited from studies conducted in mainland China, South Korea, and Japan (Supplementary Table 1). The BCAC-Asian dataset was composed of COGS (N = 10,716) and OncoArray projects (N = 14,337); twelve studies contributed samples to either or both projects. The BCAC-European dataset consisted of three sub-sets, GWAS (N = 32,498), COGS (N = 89,677), and OncoArray projects (N = 106,776)^8^.

Included as a replication set were an additional 10,829 cases and 10,996 controls of Asian ancestry, recruited by eight studies from South Korea, Japan, Hong Kong, and Taiwan (Supplementary Text 1). There was no overlap in samples from participating studies. Information on ethics approval and informed consent are described in detail for each cohort in the Supplementary Text 1.

### Genotyping and quality control

All of the genotyping and quality control procedures for GWAS, except for the expanded MEGA^EX^ chip, have been described elsewhere^3,4,6-12,34,36,37^ (Supplementary Table 1). The MEGA^EX^ chip contains approximately 2.04 million variants with an excellent genomic coverage of common variants (a minor allele frequency of 0.01 or higher) across multi-racial populations. We added to the MEGA^EX^ chip ∼80k variants selected from our GWAS of breast and colorectal cancers and exome sequencing data for breast cancer cases in Asian-ancestry populations. In total, 2.1 million variants were included on this array. We used the same quality control (QC) procedure as described elsewhere.^3,4,6-12,34,36,37^ Samples were excluded if they (i) had genotyping call rate < 95%; (ii) were male based on genotype data; (ii) had a close relationship with a Pi-HAT estimate > 0.25; (iii) were heterozygosity outliers; (iv) were ethnic outliers. SNPs were excluded if they had (i) a call rate < 95%; (ii) no clear genotyping clusters; (iii) a minor allele frequency < 0.001; (iv) a Hardy-Weinberg equilibrium test of *P* < 1 ×10^−6^; (v) genotyping concordance < 95% among the duplicated QC samples. All of the datasets were imputed using the 1000 Genomes Project Phase 3 mixed populations as the reference panel, except for the BioBank Japan (BBJ1) study, in which the HapMap Phase II (release 22) was used. Only SNPs with an imputation R^2^ > 0.3 were included in the further analyses.

Genotyping of the replication set of cases and controls was completed using the iPLEX Sequenom MassArray platform (Agena Bioscience Inc., San Diego, California, USA). One negative control (water), two blinded duplicates and two samples from the HapMap project were included as QC samples in each 96-well plate. Samples or SNPs that had a genotyping call rate of□<□95%. We also excluded SNPs that had a concordance with the QC samples of□<□95% or an unclear genotype call. If the assay could not be designed for the lead SNP, a surrogate SNP which is in LD with the lead SNP with r^2^ > 0.8 in Asians (1000 Genome) was selected. Of the 28 newly identified risk variants, 22 were successfully genotyped by Sequenom and evaluated in the association analysis, while six failed in the probe designing stage. Additional 11,642 independent samples from MYBRCA and SGBCC studies (Supplementary text 1) were also included in the replication stage in evaluation of the 22 newly identified risk variants.

### Statistical methods

Logistic regression analysis were performed within each study of Asian women to obtain a per-allele odds ratio (OR) for each SNP using PLINK2.0^38^. Principal components analyses were conducted within each GWAS dataset. Age and the top two PCs were included as covariates for in all regression models. Study (COGS) or country/region (OncoArray) was also included in the analyses of BCAC data^8^. A meta-analysis was performed using METAL^39^ with a fixed-effects model to generate Asian-specific and cross-ancestry estimates. Heterogeneity was assessed by the Cochran’s Q statistic and I^2^. For the cross-ancestry meta-analysis, we were mainly interested in evaluating variants that were associated with breast cancer risk at *P* < 0.01 in the Asian-specific analysis (n_snp_ = 244,746). However, three additional lead SNPs that did not meet this criterion can also be found in Supplementary Table 14. Inflation of the test statistics (λ) was estimated by dividing the 50th percentile of the test statistic by 0.455 (the 50th percentile for a χ^2^ distribution on 1 degree of freedom)^40^. We standardized the inflation statistic to account for the large size of our study by calculating λl,000 (λ for an equivalent study with 1,000 cases and 1,000 controls)^8^. For the replication stage, analyses were conducted with an adjustment for age and study.

Independent secondary association signals were evaluated within a flanking +/- 500kb region of the lead variant in each of the newly identified breast cancer risk loci using conditional analysis, with an adjustment for the newly identified lead risk SNPs. SNPs showing an association with breast cancer risk at *P* _conditional_ < 1 ×10^−4^ were considered independent secondary association signals. We used GCTA software (option -COJO)^41^ to perform the conditional analysis for the BBJ1, Seoul Breast Cancer Study (SeBCS), and BCAC European studies, for which only summary statistics data were available. MEGA array genotyping data was used as reference panel for LD estimation. The analysis was also performed within known susceptibility loci. All statistical tests were two-sided.

### Functional annotation

Novel risk loci were defined as those +/- 500Kb away from the lead risk variant reported by previous GWAS conducted in populations of Asian or European-ancestry for breast cancer. The lead risk SNPs newly identified in our study were defined as the variant showing an association with breast cancer risk with the lowest *P*-value in a given locus in the meta-analysis. Functional annotations of the lead SNPs and their correlated variants (r^2^ > 0.8 in 1000 Genomes Project, East Asian or European populations) were performed using HaploReg v4.1^42^. The functional annotation of chromatin states from chromHMM, DNase I hypersensitive and histone modifications such as H3K4, H3K9 and H3K27, were based on the epigenetic data in human breast mammary epithelial cells (HMEC), MCF-7 cells, and other cell lines from the Encyclopedia of DNA Elements (ENCODE) Project and Roadmap Epigenetics Project.

### Expression quantitative loci (eQTL) analysis

To identify target genes, we performed eQTL analysis utilized four independent sets of gene expression data derived from normal breast (N= 85, GTEx, women of European ancestry), breast tumor (women of European ancestry, TCGA, N = 672; METABRIC, N = 1,904) and adjacent normal tissues (women of Asian ancestry, SBCGS, N = 151). We focused on *cis-*eQTL analyses for genes residing ± 500 Kb flanking each newly associated leading SNP. The details of data processing were described in Supplementary Text 2.

A linear regression model was used to perform eQTL analyses to estimate the additive effect for each leading SNP identified on gene expression levels. We additionally adjusted for somatic copy number alteration and methylation levels in the regression model for the analysis of TCGA data. We only adjusted for somatic copy number alteration in the analysis for the METABRIC set.

### Gene-based analysis

We recently conducted a transcriptome wide association study (TWAS) to investigate associations of genetically predicted gene expression with the risk of breast cancer^43^. We utilized the same approach to examine the associations with breast cancer risk of genes located within flanking 500kb of each newly associated leading SNP. The breast-specific prediction model was generated using the elastic net method as implemented in the glmnet R package (α= 0.5), with tenfold cross-validation^43^. To further increase statistical power, we also utilized 6,124 samples across 39 tissue types from 369 unique European individuals who had genome-wide genotype data available to build cross-tissue models, as previously described^44,45^. The expression of a gene for individual *i* in tissue *t, Y*_*i,t*_, is modeled as 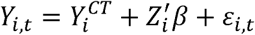, where 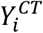 represents the cross-tissue component of expression levels for a given gene. The mixed effect model parameters were estimated using the lme4 package in R. The predicted gene expressions 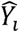 in the breast-specific models and 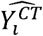 in the cross-tissue models then were evaluated for their associations with breast cancer risk in the ABCC and BCAC, using methods implemented in MetaXcan^46^.

## Data Availability

The data are available through contacting BCAC and ABCC consortia.

## REFERENCES

1. Torre, L.A. et al. Global cancer statistics, 2012. CA Cancer J Clin 65, 87–108 (2015).

2. Shiovitz, S. & Korde, L.A. Genetics of breast cancer: a topic in evolution. Ann Oncol 26, 1291–9 (2015).

3. Zheng, W. et al. Genome-wide association study identifies a new breast cancer susceptibility locus at 6q25.1. Nat Genet 41, 324–8 (2009).

4. Cai, Q. et al. Genome-wide association analysis in East Asians identifies breast cancer susceptibility loci at 1q32.1, 5q14.3 and 15q26.1. Nat Genet 46, 886–90 (2014).

5. Wellcome Trust Case Control, C. Genome-wide association study of 14,000 cases of seven common diseases and 3,000 shared controls. Nature 447, 661–78 (2007).

6. Zheng, W. et al. Common genetic determinants of breast-cancer risk in East Asian women: a collaborative study of 23 637 breast cancer cases and 25 579 controls. Hum Mol Genet 22, 2539–50 (2013).

7. Michailidou, K. et al. Genome-wide association analysis of more than 120,000 individuals identifies 15 new susceptibility loci for breast cancer. Nat Genet 47, 373–80 (2015).

8. Michailidou, K. et al. Association analysis identifies 65 new breast cancer risk loci. Nature 551, 92–94 (2017).

9. Cai, Q. et al. Genome-wide association study identifies breast cancer risk variant at 10q21.2: results from the Asia Breast Cancer Consortium. Hum Mol Genet 20, 4991–9 (2011).

10. Long, J. et al. Genome-wide association study in east Asians identifies novel susceptibility loci for breast cancer. PLoS Genet 8, e1002532 (2012).

11. Long, J. et al. A common deletion in the APOBEC3 genes and breast cancer risk. J Natl Cancer Inst 105, 573–9 (2013).

12. Han, M.R. et al. Genome-wide association study in East Asians identifies two novel breast cancer susceptibility loci. Hum Mol Genet 25, 3361–3371 (2016).

13. Jia, W.H. et al. Genome-wide association analyses in East Asians identify new susceptibility loci for colorectal cancer. Nat Genet 45, 191–6 (2013).

14. Zhang, B. et al. Large-scale genetic study in East Asians identifies six new loci associated with colorectal cancer risk. Nat Genet 46, 533–42 (2014).

15. Zeng, C. et al. Identification of Susceptibility Loci and Genes for Colorectal Cancer Risk. Gastroenterology 150, 1633–1645 (2016).

16. Lu, Y. et al. Large-Scale Genome-Wide Association Study of East Asians Identifies Loci Associated With Risk for Colorectal Cancer. Gastroenterology 156, 1455–1466 (2019).

17. Kalitsis, P., Zhang, T., Marshall, K.M., Nielsen, C.F. & Hudson, D.F. Condensin, master organizer of the genome. Chromosome Res 25, 61–76 (2017).

18. Wang, H.Z., Yang, S.H., Li, G.Y. & Cao, X. Subunits of human condensins are potential therapeutic targets for cancers. Cell Div 13, 2 (2018).

19. Krepischi, A.C. et al. Germline DNA copy number variation in familial and early-onset breast cancer. Breast Cancer Res 14, R24 (2012).

20. Horpaopan, S. et al. Genome-wide CNV analysis in 221 unrelated patients and targeted high-throughput sequencing reveal novel causative candidate genes for colorectal adenomatous polyposis. Int J Cancer 136, E578–89 (2015).

21. He, L. et al. alpha-Mannosidase 2C1 attenuates PTEN function in prostate cancer cells. Nat Commun 2, 307 (2011).

22. Schulz, E. et al. Germline variants in the SEMA4A gene predispose to familial colorectal cancer type X. Nat Commun 5, 5191 (2014).

23. Chang, J. et al. Pre-clinical evaluation of small molecule LOXL2 inhibitors in breast cancer. Oncotarget 8, 26066–26078 (2017).

24. Chang, A.C. et al. STC1 expression is associated with tumor growth and metastasis in breast cancer. Clin Exp Metastasis 32, 15–27 (2015).

25. Lindstrom, S. et al. Genome-wide association study identifies multiple loci associated with both mammographic density and breast cancer risk. Nat Commun 5, 5303 (2014).

26. McCormack, V.A. & dos Santos Silva, I. Breast density and parenchymal patterns as markers of breast cancer risk: a meta-analysis. Cancer Epidemiol Biomarkers Prev 15, 1159–69 (2006).

27. Al Olama, A.A. et al. A meta-analysis of 87,040 individuals identifies 23 new susceptibility loci for prostate cancer. Nat Genet 46, 1103–9 (2014).

28. Litchfield, K. et al. Identification of 19 new risk loci and potential regulatory mechanisms influencing susceptibility to testicular germ cell tumor. Nat Genet 49, 1133–1140 (2017).

29. Rafnar, T. et al. Variants associating with uterine leiomyoma highlight genetic background shared by various cancers and hormone-related traits. Nat Commun 9, 3636 (2018).

30. Migliorini, G. et al. Variation at 10p12.2 and 10p14 influences risk of childhood B-cell acute lymphoblastic leukemia and phenotype. Blood 122, 3298–307 (2013).

31. O’Mara, T.A. et al. Identification of nine new susceptibility loci for endometrial cancer. Nat Commun 9, 3166 (2018).

32. McKay, J.D. et al. Large-scale association analysis identifies new lung cancer susceptibility loci and heterogeneity in genetic susceptibility across histological subtypes. Nat Genet 49, 1126–1132 (2017).

33. Diskin, S.J. et al. Common variation at 6q16 within HACE1 and LIN28B influences susceptibility to neuroblastoma. Nat Genet 44, 1126–30 (2012).

34. Elgazzar, S. et al. A genome-wide association study identifies a genetic variant in the SIAH2 locus associated with hormonal receptor-positive breast cancer in Japanese. J Hum Genet 57, 766–71 (2012).

35. Xiao, R. & Boehnke, M. Quantifying and correcting for the winner’s curse in genetic association studies. Genet Epidemiol 33, 453–62 (2009).

36. Zhang, Y. et al. Rare coding variants and breast cancer risk: evaluation of susceptibility Loci identified in genome-wide association studies. Cancer Epidemiol Biomarkers Prev 23, 622–8 (2014).

37. Kim, H.C. et al. A genome-wide association study identifies a breast cancer risk variant in ERBB4 at 2q34: results from the Seoul Breast Cancer Study. Breast Cancer Res 14, R56 (2012).

38. Chang, C.C. et al. Second-generation PLINK: rising to the challenge of larger and richer datasets. Gigascience 4, 7 (2015).

39. Willer, C.J., Li, Y. & Abecasis, G.R. METAL: fast and efficient meta-analysis of genomewide association scans. Bioinformatics 26, 2190–1 (2010).

40. Devlin, B. & Roeder, K. Genomic control for association studies. Biometrics 55, 997–1004 (1999).

41. Yang, J., Lee, S.H., Goddard, M.E. & Visscher, P.M. GCTA: a tool for genome-wide complex trait analysis. Am J Hum Genet 88, 76–82 (2011).

42. Ward, L.D. & Kellis, M. HaploReg v4: systematic mining of putative causal variants, cell types, regulators and target genes for human complex traits and disease. Nucleic Acids Res 44, D877–81 (2016).

43. Wu, L. et al. A transcriptome-wide association study of 229,000 women identifies new candidate susceptibility genes for breast cancer. Nat Genet 50, 968–978 (2018).

44. Wheeler, H.E. et al. Survey of the Heritability and Sparse Architecture of Gene Expression Traits across Human Tissues. PLoS Genet 12, e1006423 (2016).

45. Lu, Y. et al. A Transcriptome-Wide Association Study Among 97,898 Women to Identify Candidate Susceptibility Genes for Epithelial Ovarian Cancer Risk. Cancer Res 78, 5419–5430 (2018).

46. Barbeira, A.N. et al. Exploring the phenotypic consequences of tissue specific gene expression variation inferred from GWAS summary statistics. Nat Commun 9, 1825 (2018).

